# Cardiovascular Disease Risk Factor Control Following Release from Carceral Facilities

**DOI:** 10.1101/2024.03.14.24304323

**Authors:** JA Aminawung, LB Puglisi, B. Roy, N Horton, JE Elumn, H. Lin, K Bibbins-Domingo, H. Krumholz, EA Wang

## Abstract

**Background:** Incarceration is a social determinant of cardiovascular health but is rarely addressed in clinical settings or public health prevention efforts. People who have been incarcerated are more likely to develop cardiovascular disease (CVD) at younger ages and have worse cardiovascular outcomes compared with the general population, even after controlling for traditional risk factors. This study aims to identify incarceration-specific factors that are associated with uncontrolled CVD risk factors to identify potential targets for prevention.

**Methods:** Using data from Justice-Involved Individuals Cardiovascular Disease Epidemiology (JUSTICE), a prospective cohort study of individuals released from incarceration with CVD risk factors, we examine the unique association between incarceration-specific factors and CVD risk factor control, including systolic blood pressure (SBP≥140 mmHg, diastolic blood pressure (DBP)≥90, body mass index (BMI)≥40, glycosylated hemoglobin (HbA1c) ≥8%, and low-density lipoprotein cholesterol (LDL-c)≥ 160). Incarceration-specific factors include the conditions of confinement (jail vs. prison, time in solitary confinement), and collateral sanctions following release (barriers to housing, food, employment due to criminal record). Variables associated with uncontrolled CVD risk factors were included in the multivariate model to examine the unique contribution of each risk factor with uncontrolled CVD risk factors.

**Results:** Participants (N=471), mean age of 45.0 ±SD 10.8 years were disproportionately men (89%), from racially minoritized groups (79%), and poor (91% below the 100% federal poverty level). Over half (54%) had at least one uncontrolled CVD risk factor at baseline. People released from jail, compared with prison, had lower Life’s Essential 8 scores for blood pressure and smoking. Having been incarcerated in jail, as compared with prison, was associated with an increased odds of having an uncontrolled CVD risk factor, even after adjusting for age, race and ethnicity, gender, perceived stress, and life adversity score (AOR 1.62, 95% CI 1.02-2.57).

**Discussion:** Release from jail is associated with poor CVD risk factor control and requires tailored intervention, which is informative as states design and implement the Centers of Medicare & Medicaid Services Reentry 1115 waiver, which allows Medicaid to cover services prior to release from correctional facilities.

## Introduction

Mass incarceration and its effects on cardiovascular health remains understudied and largely invisible to the scientific community despite its effect on the 80 million individuals who have a criminal record and their families and communities. The term mass incarceration refers to the unparalleled rates of incarceration in the United States (US), its disproportionate concentration among Black families and communities, and the vast system of laws that maintain this status quo and prevent people with criminal records from obtaining publicly-funded services and from re-entering the labor force.^1^ There are 1.9 million individuals currently incarcerated and over 7 million incarcerations yearly, disproportionately of racially minoritized populations.^2^ Recent data from the Bureau of Justice Statistics show cardiovascular disease (CVD) to be a leading cause of death among people incarcerated in state prisons and local jails,^3^ where the imprisonment of Black men is six times higher and Black women double the rates of their White counterparts.^4,5^ Following release from incarceration, people have higher hospitalization rates from complications of diabetes and hypertension and increased CVD mortality compared with the general population.^6,7^

The impact of incarceration on CVD risk traverses the life course, as incarceration in young adulthood is associated with incident hypertension and the development of left ventricular hypertrophy and CVD 20 to 30 years after the imprisonment episode, even after controlling for clinical risk factors and sociodemographic characteristics.^8,9^ Incarceration may impact CVD risk factor management in various ways unique to the carceral experience. Reduced access to illicit drugs and alcohol, a ban on smoking in almost all federal prisons and a majority of state prisons,^10^ and enforced medication adherence may improve overall health while incarcerated.^11^ But individuals have limited opportunities for exercise, choice of diet, and self-management of medications while imprisoned, especially in the most extreme conditions in maximum security prisons or solitary confinement, where movement is extremely restricted as people are detained in their cells for up to 23 hours a day.^12,13^

Release from carceral systems can introduce further precarity, as many prisons and jails do not provide discharge planning, including medications or referrals to primary care or specialty appointments.^14^ Many individuals have had limited to no use of the community health care system prior to incarceration and do not know how to obtain their medications from a pharmacy or self-manage their CVD risk factors. Close to two thirds of people with health conditions released from incarceration have been found to have inadequate health literacy, which is associated with decreased confidence in taking medications and increased emergency department visits in the community.^15^ After release, the collateral sanctions of having a criminal record or being surveilled by community corrections through parole or probation can further affect their health.^16^ The National Inventory of Collateral Consequences of Conviction notes 44,000 laws that prohibit the provision of social needs and civil rights of people who have criminal records, including lifetime voter disenfranchisement, housing restrictions, food stamp bans, barriers to employment and healthcare access, and restitutions of debt to the criminal legal system.^17^ These health-related social needs directly impact cardiovascular disease risk factor prevalence and control.^18^

Almost a decade ago, the National Heart, Lung, Blood Institute (NHLBI) and American Heart Association summarized in separate reports the need to better understand the unique impact of mass incarceration on cardiovascular health.^19,20^ They highlighted the importance of studying how mass incarceration affects CVD risk factor control immediately following release and conceptualized incarceration’s impact as a place of residence and a system of practices and policies that affect carceral healthcare delivery. But this is challenging as national health surveillance studies and existing NHLBI cohort studies do not include detailed measures of individual level exposure to mass incarceration.^20,21^ We aimed to describe how mass incarceration affects CVD risk factor control among individuals just released from carceral facilities, hypothesizing that the conditions of one’s confinement and collateral sanctions following release would affect CVD risk factor control. By identifying various features of mass incarceration that affect CVD risk factors, we can identify novel place-based and legal and policy prevention efforts to improve CVD risk factor control for the millions of Americans impacted by incarceration.

## Methods

### Study Design

This study uses data from a prospective, observational cohort, **JUST**ice-Involved **I**ndividuals **C**ardiovascular Disease **E**pidemiology (JUSTICE, R01 HL137696), whose protocol has been described in detail previously.^22^ Briefly, we recruited individuals released from a carceral facility to New Haven, Bridgeport, and Hartford, Connecticut. Connecticut is one of six states in the country with a unified prison and jail system (prisons house people incarcerated for sentences longer than one year, and jails house those awaiting adjudication of crime or people with sentences of less than a year, in general) and has one of the highest racial disparities in sentencing in the country (over 70% of the incarcerated population are Black and Latino individuals).^23^ These features make it an ideal setting to study how differential exposure to carceral systems affects CVD risk factors, primarily among racially minoritized populations.

### Population

Eligibility criteria for the JUSTICE study included release from a carceral facility within the past three months and the presence of any of the following CVD risk factors: hypertension, diabetes, obesity, or hyperlipidemia. We chose to include participants with baseline CVD risk factors to target individuals at higher risk of poor CVD outcomes, improving our ability to measure the impact of criminal legal exposures during incarceration and in the community on CVD risk factor control. We excluded individuals with a known terminal illness, given the intention to follow participants for 12 months, and with serious mental illness, as they may be limited in their ability to consent to study participation.

### Study Procedures

At the baseline visit, we used a teach-to-goal method developed to ensure participant understanding of the study design and protocol.^24^ All participants provided informed written consent. A trained research assistant administered a baseline survey, gathering information on sociodemographic data, clinical, psychosocial, behavioral, and medical factors associated with CVD risk factors, and exposure to incarceration and community corrections policies related to CVD management and collateral sanctions (barriers to employment, housing, and food based on criminal record) following release. We also measured participant weight, height, and blood pressure and then performed point-of-care testing to measure lipids and glycosylated hemoglobin. We measured participant blood pressure in a sitting position after they had rested for at least 15 minutes, with a second reading taken, if necessary, per study protocol, using an Omron Hem 705CP^®^ blood pressure monitor and an appropriately sized cuff.^25^ We measured participants’ weight using a digital medical grade floor scale and height using a stadiometer and computed body mass index (kg/m^2^) from the participant’s weight and height values. We obtained total cholesterol and high-density lipoprotein cholesterol (HDL-c) concentration using a portable Cardiocheck^®^ professional analyzer and HbA1c using A1CNow+^®^ multi-test point of care testing system.^26^ The visit activities lasted between 60 and 90 minutes on average. Midway through this study, we were awarded a supplement that enabled us to measure sleep health, as previously described.^27^ All participants were compensated $60 for their time to complete the baseline study procedures.

### Criminal legal exposure

We examined exposure to incarceration as a place of residence and a system of laws and practices that affect individuals during imprisonment and following release as independent variables of interest. We asked participants the total number of times and the total number of years they had been incarcerated over their lifetime and computed the most recent incarceration length from self-reported incarceration start and end dates. We inquired where they served their most recent (index) incarceration (jail/prison) and exposure to solitary confinement (also referred to as residential segregation or administrative segregation), a carceral system sanction in which people are held in 8 by 6-foot cells for close to 23 hours daily. Participants’ residence following release was categorized by whether they resided in a halfway house (a transitional house where they remain under the carceral system, where people have less opportunity to control their diet and exercise) or lived in other congregate settings or home.

Participants were also asked if they were experiencing post-incarceration collateral sanctions, including difficulties obtaining employment, driver’s and occupational licenses, government benefits, housing, food access due to a criminal record, and wage garnishment for child support and other forms of debt. We also asked whether they had conditions to their release, including monitoring through parole (early release mechanism from prison) and probation (supervision of people in the community in lieu of incarceration after conviction.)

### Cardiovascular disease risk factor control

The primary dependent variable was any uncontrolled CVD risk factor, which was defined as systolic blood pressure (SBP )≥140 mmHg or diastolic blood pressure (DBP) ≥ 90 mmHg, body mass index (BMI) ≥40 kg/m^2^, glycosylated hemoglobin (HbA1c) ≥8%, or low-density lipoprotein cholesterol (LDL-c)≥ 160), based on the values measured at the baseline visit.^28,29^

### Cardiovascular Health

We measured cardiovascular health using the American Heart Association’s Life’s Essential Eight (LE8) to identify areas for possible prevention and because cardiovascular health is associated with mortality.^30^ The LE8 score (0–100 scale) was the average score for eight individual components (blood pressure, cholesterol, blood glucose, body mass index, diet, physical activity, sleep, and smoking) that affect cardiovascular health. In the general population, an average score of 0 to 49 is defined as *low* cardiovascular health, 50 to 79 is *moderate* cardiovascular health, and 80 to 100 is *high* cardiovascular health.^31^ Below we describe how we scored each of the eight components on a 0-100 scale.

#### Diet

Diet was assessed using an adapted version of the Eating At America’s Table Study (EATS) questionnaire.^32^ Participants reported how often they ate a serving of vegetables, fruits, or foods cooked in fat since being released from prison. After reversing the foods cooked in the fat score, we added the scores from each food to obtain the EATS score. Based on an NIH “all-day” diet screener score, we categorized the scores as follows: a score of less than 4 received 0 points.^33^ Those who scored between 4 and 8 received between 25 and 75 points, and those with a score of 8 and above received 100 points.

#### Physical Activity

Participants were asked if they participated in any vigorous activities outside of work.^34^ To obtain physical activity time, we multiplied the number of times per week by the exercise duration in minutes. Individuals who reported not participating in vigorous activity outside of work were given 0 points. Greater than or equal to 150 physical activity minutes received 100 points. Physical activity minutes between 0 and 150 minutes and received scores from 20 to 90, as described in Table 1.

**Table 1:** Measurement and Scoring of Life’s Essential 8. ^31^

| Cardiovascular health metric | Method of measurement | Scoring |  |
| --- | --- | --- | --- |
| Diet | Eating at America's Table (EATS)<br>"All Day Screener" | **Points** | **EATS score** |
|  |  | 100 | 8.0 – 10 |
|  |  | 80 | 6.0 – 7.99 |
|  |  | 50 | 5.0 – 5.99 |
|  |  | 25 | 4.0 – 4.99 |
|  |  | 0 | 0 – 3.99 |
| Physical activity | Physical activity questionnaire,<br>min/week | **Points** | **Minutes/week** |
|  |  | 100 | ≥150 |
|  |  | 90 | 120–149 |
|  |  | 80 | 90–119 |
|  |  | 60 | 60–89 |
|  |  | 40 | 30–59 |
|  |  | 20 | 1–29 |
|  |  | 0 | 0 |
| Smoking | Smoking history | **Points** | **Status** |
|  |  | 100 | Never smoker |
|  |  | 75 | Quit (other) |
|  |  | 50 | Quit (≥ 5 cig/week before last incarceration) |
|  |  | 25 | Quit (≥ 100 cigarettes in lifetime) |
|  |  | 0 | Current smoker |
| Sleep | Average sleep per night | **Points** | **Hours of sleep** |
|  |  | 100 | 7 to <9 |
|  |  | 90 | 9 to <10 |
|  |  | 70 | 6 to <7 |
|  |  | 40 | 5 to <6 or ≥10 |
|  |  | 20 | 4 to <5 |
|  |  | 0 | <4 |
| Body mass index | (Weight (kg)/height (m)<sup>2</sup>) | **Points** | **BMI, kg/m<sup>2</sup>** |
|  |  | 100 | <25 |
|  |  | 70 | 25.0–29.9 |
|  |  | 30 | 30.0–34.9 |
|  |  | 15 | 35.0–39.9 |
|  |  | 0 | ≥40.0 |
| Non-HDL-c | Point of care measured total cholesterol minus HDL-c | **Points** | **Non-HDL-c, mg/dL** |
|  |  | 100 | <130 |
|  |  | 60 | 130–159 |
|  |  | 40 | 160–189 |
|  |  | 20 | 190–219 |
|  |  | 0 | ≥220 |
| Blood glucose | Glycosylated hemoglobin | **Points** | **%** |
|  |  | 100 | <5.7 |
|  |  | 60 | 5.7–6.3 |
|  |  | 40 | 6.4 – 6.9 |
|  |  | 30 | 7.0–7.9 |
|  |  | 20 | 8.0–8.9 |
|  |  | 10 | 9.0–9.9 |
|  |  | 0 | ≥10.0 |
| Blood pressure | Measured systolic and diastolic blood pressure, mm Hg | **Points** | **mm Hg** |
|  |  | 100 | <120/<80 |
|  |  | 75 | 120–129/<80 |
|  |  | 50 | 130–139 or 80–89 |
|  |  | 25 | 140–159 or 90–99 |
|  |  | 0 | ≥160 or ≥100 |

#### Sleep

Participants’ average hours of sleep since release from a carceral facility were assessed using the Pittsburg Sleep Quality Index (PSQI).^35^ The optimal sleep was 7 to <9 hours per night (100 points), and 0 points were awarded for sleep <4 hours per night. Other sleep durations received points between 0 and 100, as described in Table 1. The first 154 participants had missing data on PSQI because they we enrolled before the sleep supplement was awarded. For these participants, we employed a regression-based imputation on participant age, gender, place of residence, perceived stress, and self-efficacy.

#### Smoking

Smoking status was self-reported through a series of questions that assessed participants’ use of cigarettes. Because of restricted access to cigarettes while incarcerated or even after release, if they were living in a halfway house, we ascertained their smoking history with several questions. We asked about lifetime smoking history, including if they had smoked more than 100 cigarettes in their lifetime, smoked regularly before the last incarceration, and if they currently smoked cigarettes. Current smokers received 0 points. Participants who were no longer smoking received between 25 and 75 points, depending on lifetime cigarette use, as described in Table 1. Participants who never smoked received 100 points.

#### Body Mass Index

A body mass index score of less than 25 kg/m^2^ received 100 points, while a score of 40 kg/m^2^ and above received 0 points. Scores between 25 and 40 kg/m^2^ were assigned points between 15 and 70.

#### Cholesterol

Non-HDL-c (total cholesterol minus HDL-c) of <130 mg/dL was considered optimal (100 points). Non-HDL-c of 220 mg/dL or more received 0 points. Values between 130 and 220 mg/dL received scores between 20 and 60.

#### Blood Glucose

A HbA1c of <5.7% received 100 points. Participants with an HbA1c ≥10.0% received 0 points. Values of HbA1c between 5.7% and 10.0% received scores between 10 and 60.

#### Blood Pressure

Participants with a systolic blood pressure of <120 and diastolic blood pressure of <80 mmHg received 100 points. Participants with a systolic blood pressure of ≥160 mmHg and diastolic blood pressure of ≥100 mmHg received zero points. Systolic blood pressure values between 120 and 159mmHg and diastolic values between 80 and 99mmHg, were scored between 25 and 75 points.

### Demographic Variables (Covariates)

The participant’s age at the baseline visit was calculated in years from the participant’s self-reported date of birth. Gender was self-reported at the time of the interview with male, female, and transgender being the only options for selection. Race and ethnicity were self-reported from the options non-Hispanic White, non-Hispanic Black, non-Hispanic Other (including Asian, Hawaiian, and Native American), and Hispanic. We conceptualized race and ethnicity as social constructs that result in differential exposure to incarceration and health care due to structural racism.^36^ We include measures of race and ethnicity in these analyses to highlight the disproportionate impact of mass incarceration on those who are racialized as Black individuals and to determine whether the observed association between carceral exposures and CVD risk factor control is different among those of a minoritized population.

### Analytical approach

We described participants characteristics and then, using t-tests, Mann Whitney U test and chi square tests, we examined CVD risk factor control at baseline by sociodemographic and clinical characteristics, exposure to various features of mass incarceration, especially conditions of confinement and collateral consequences of having a criminal record. We next examined factors associated with having any uncontrolled CVD risk factor using multivariable logistic regression by adjusting for all the covariates related to uncontrolled CVD risk factors in the bivariate analyses. We computed component and total LE8 scores, and by place of last incarceration, to identify targets for policy and prevention efforts to reduce the carceral systems’ impact on cardiovascular health. A sensitivity analysis excluding participants with imputed sleep scores yielded similar sleep component and total LE8 scores. Thus, we elected to report scores for the whole sample. Statistical analyses were performed in SPSS version 29.0.1. Statistical significance for all analyses was defined as 2-sided alpha =0.05. The Yale Human Investigation Committee and the Office of Human Research Protections approved this study.

## Results

The mean age of participants (N=471) was 45.0 years ± standard deviation (SD) 10.8 and ranged from 22 to 71 years. Participants were disproportionately men (89%), of racially minoritized groups (79%), and poor (91% below the 100% federal poverty level), reflective of the Connecticut Department of Correction population. Median incarceration length of those returning from jail was 9 months and 26 months for those returning from prison. Over three-quarters of participants (77%) lived in halfway houses, 20% reported going 24 hours without food in the past month, and 86% reported being unemployed. Seventy-eight percent of participants had been diagnosed with hypertension, 47% with hyperlipidemia, and 28% with diabetes. (Table 2).

**Table 2.** Participant characteristics at baseline visit (N=471).

| **Sociodemographic characteristic** | **No.** | **%** |
| --- | --- | --- |
| Age, mean (SD), [range] | 45.0 (10.8), [21-72] |  |
| Race |  |  |
| Hispanic | 128 | 27.2% |
| Non-Hispanic Black | 229 | 48.6% |
| Non-Hispanic Other | 13 | 2.8% |
| Non-Hispanic White | 101 | 21.4% |
| Gender |  |  |
| Female | 51 | 10.8% |
| Male | 420 | 89.2% |
| Education |  |  |
| Less than high school | 109 | 23.1% |
| GED | 97 | 20.6% |
| High school graduate | 138 | 29.3% |
| Some college/technical education | 112 | 23.8% |
| College graduate | 15 | 3.2% |
| Marital status |  |  |
| Single | 355 | 75.4% |
| Married | 44 | 9.3% |
| Separated, widowed divorced | 72 | 15.3% |
| Average monthly income |  |  |
| No income | 309 | 65.6% |
| Less than \$500 | 92 | 19.5% |
| \$500-\$999 | 21 | 4.5% |
| \$1000 or more | 43 | 9.1% |
| **Clinical Factors (history)** |  |  |
| Diabetes | 132 | 28.0% |
| Coronary Disease | 63 | 13.4% |
| Hypercholesteremia | 219 | 46.5% |
| Hypertension | 369 | 78.3% |
| Obesity, BMI≥30 kg/m<sup>2</sup> | 283 | 60.1% |
| History of drug use disorder | 273 | 58.0% |
| History of alcohol use disorder | 124 | 26.3% |
| **Behavioral Factors (post-release)** |  |  |
| Smoking | 277 | 58.8% |
| Number of current daily cigarettes*, median (IQR) | 5 (6) |  |
| Alcohol Use | 20 | 4.2% |
| Cocaine Use | 18 | 3.8% |
| Physical exercise | 248 | 53% |
| **Psychosocial factors, mean (SD), [range]** |  |  |
| Everyday Discrimination (9-items) | 15.41 (7.10), [9-47] |  |
| Perceived Stress Scale (10-items) | 15.84 (7.29), [0-38] |  |
| General Self-Efficacy Scale (10-items) | 33.06 (5.33), [1-40] |  |
| Depression (PHQ-9) (9-items) | 7.10 (6.01), [0-26] |  |
| PTSD Symptom Scale (17-items) | 20.63 (14.21), [0-51] |  |
| Cumulative Adversity Interview (33-items) | 16.77 (6.68), [0-31] |  |
| **Structural factors** |  |  |
| Employed | 67 | 14.2% |
| Health Insurance | 440 | 93.4% |
| Insurance Type |  |  |
| Medicaid | 423 | 96.1% |
| Medicare | 14 | 3.2% |
| Have a primary care provider | 217 | 46.1% |
| Housing Status |  |  |
| Homeless | 19 | 4.0% |
| Transitional Home | 368 | 78.1% |
| Living with Family/Friends | 43 | 9.1% |
| Rent or Own | 41 | 8.7% |
| Living in a halfway house | 363 | 77.1% |
| **Correctional specific factors/collateral sanctions** |  |  |
| Lifetime incarceration |  |  |
| One time | 50 | 10.6% |
| 2 - 5 times | 205 | 43.5% |
| 6-10 times | 99 | 21.0% |
| More than 10 times | 113 | 24.0% |
| Lifetime incarcerated; months, median (IQR) | 132 (180) |  |
| Index incarceration |  |  |
| Jail | 103 | 21.9% |
| Prison | 368 | 78.1% |
| Length of incarceration; months, median (IQR) | 21 (42) |  |
| On Parole | 222 | 47.1% |
| On Probation | 97 | 20.6% |
| Solitary confinement during last Incarceration | 178 | 37.8% |
| Saw health care provider during last incarceration | 335 | 71.1% |
| Co-payment when incarcerated† | 196 | 58.5% |
| Discriminated by health care providers† | 192 | 57.3% |
| Receiving disability insurance | 19 | 4.0% |
| Receiving Food Stamps | 126 | 26.8% |
| Without food for a whole day since release | 94 | 20.0% |
| Child Support | 91 | 19.3% |
| Barriers to Employment / Licensure based on criminal record | 348 | 73.9% |
| Debt / Restitution | 170 | 36.1% |
| Barriers to Housing based on criminal record | 39 | 8.3% |

Notes: SD=standard deviation, IQR= interquartile range; PTSD = Post traumatic stress disorder; PHQ= Patient health questionnaire
\*=current smokers; † = participants who reported seeing a provider during last incarceration;
††=participants who reported taking medications.

Over half of the participants (N=252) had at least one uncontrolled CVD risk factor at baseline. Those with uncontrolled CVD risk factors were older compared to those with controlled CVD risk factors (45.9 vs. 43.9 years). We observed no differences in CVD risk factor control by gender or self-reported race and ethnicity (Table 3). There were also no differences in CVD risk factor control by housing status, marital status, or any measure of socioeconomic status, including educational attainment, employment status, current income, or insurance status.

**Table 3:**
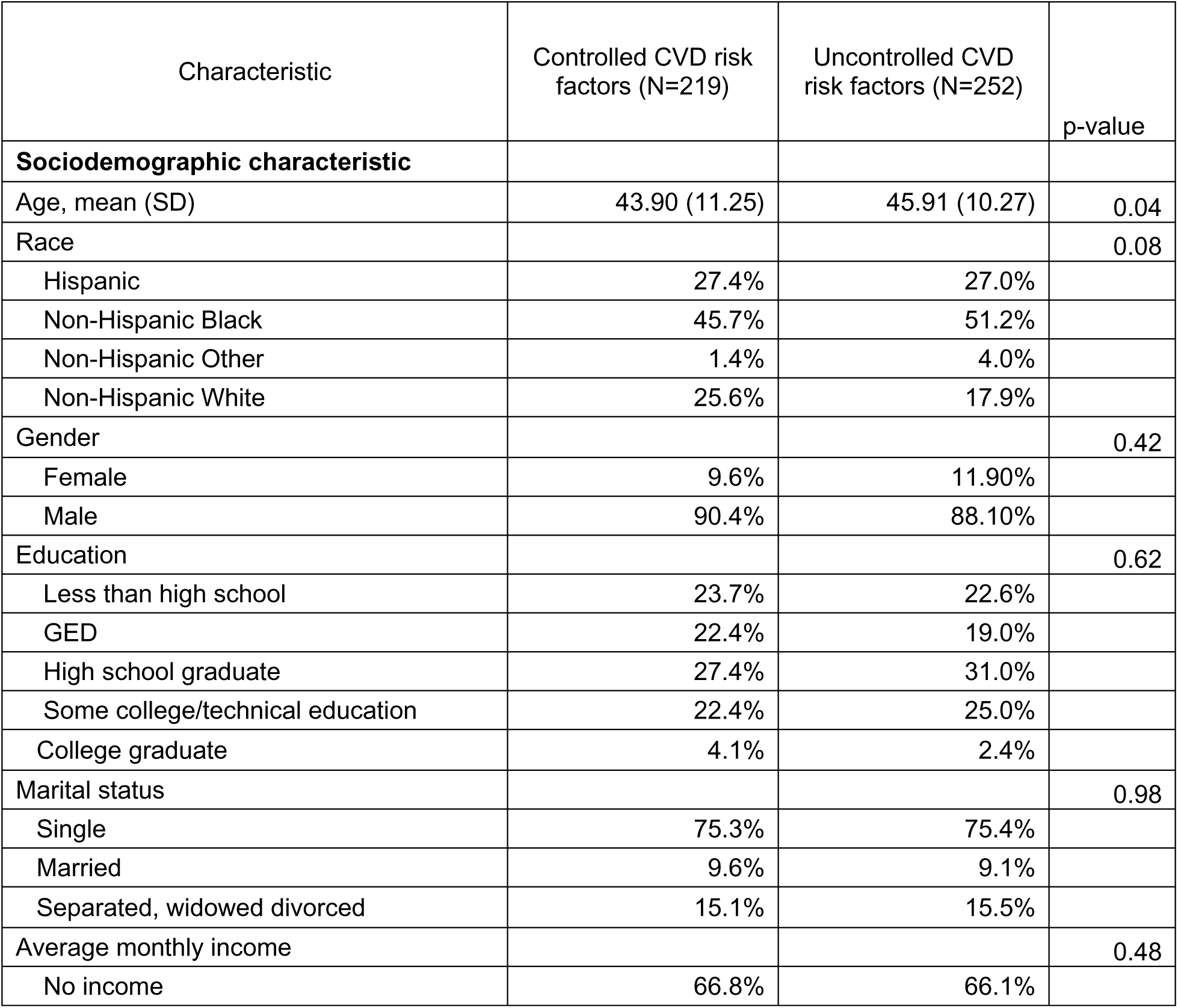
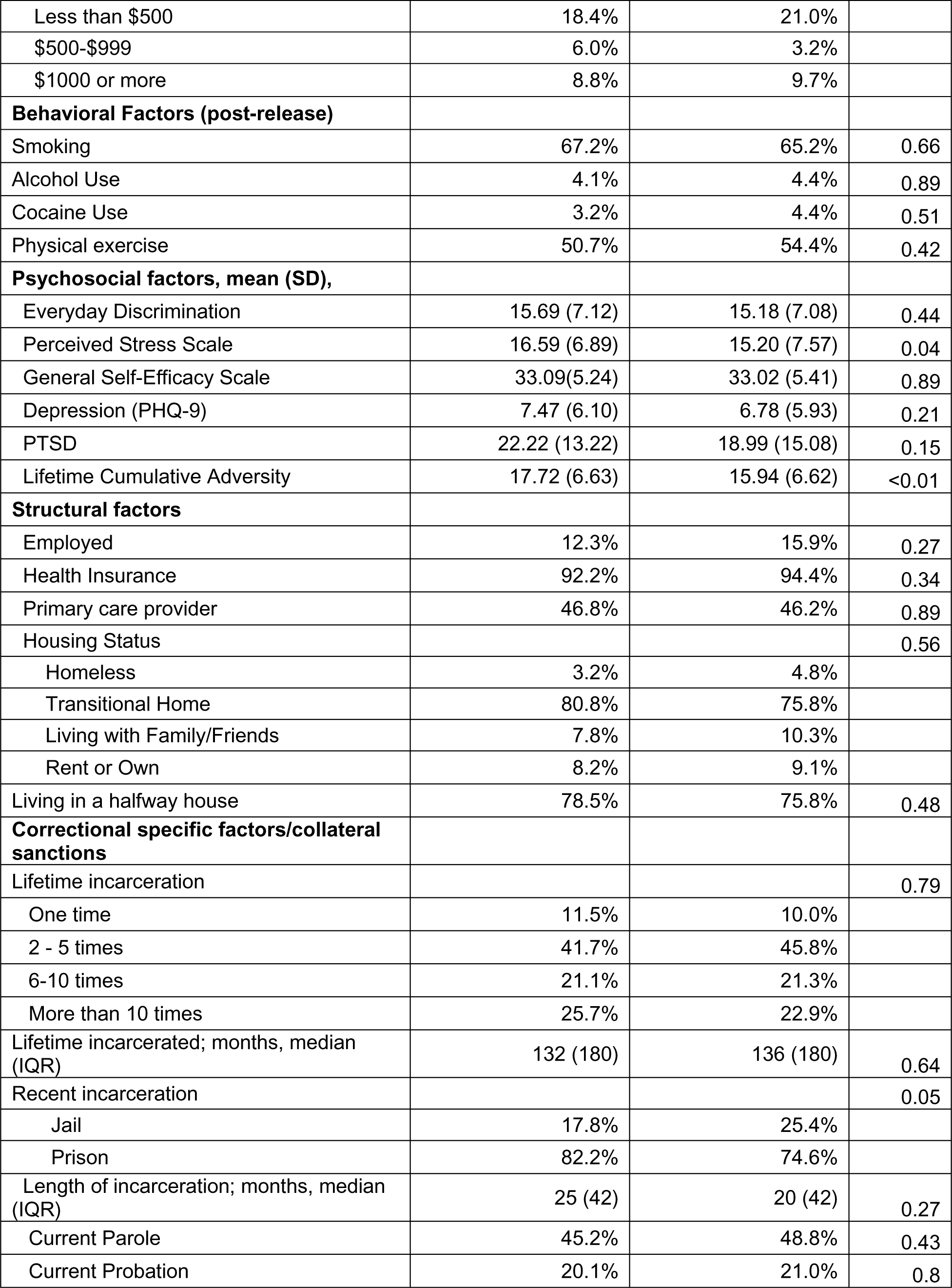
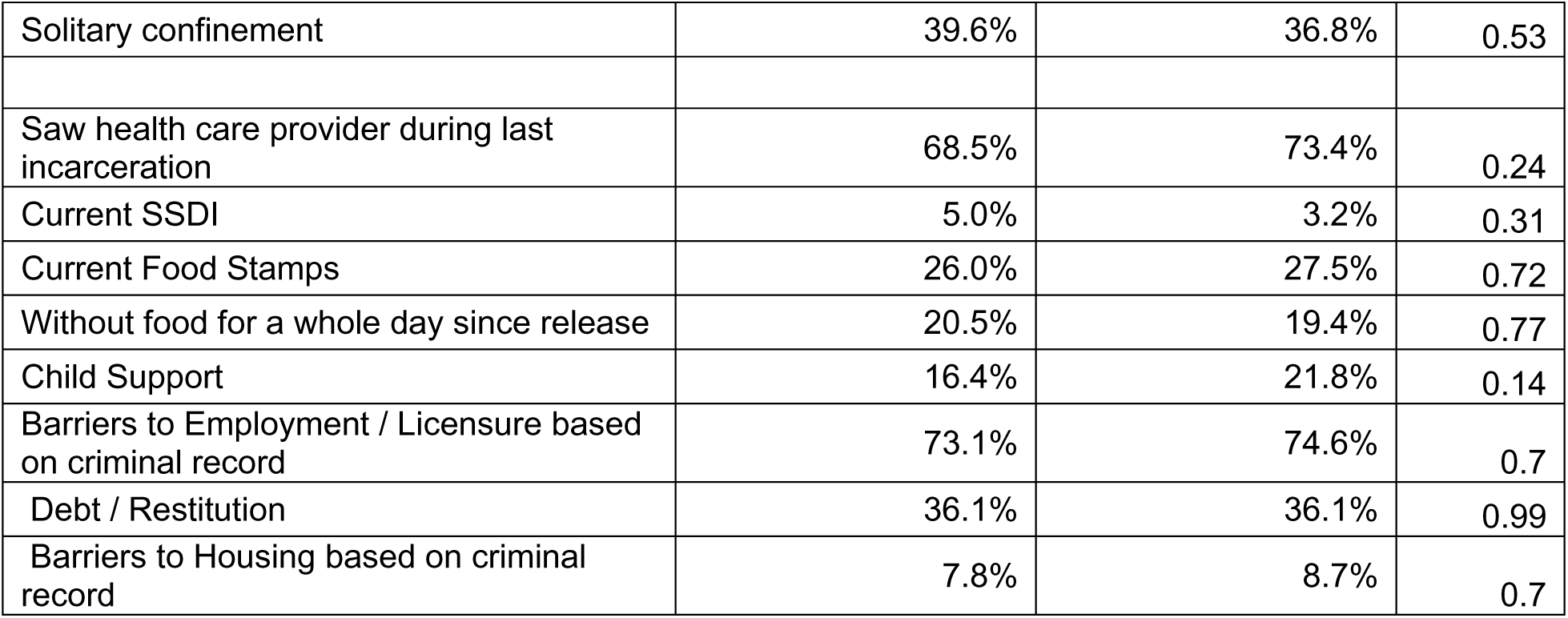
Participant characteristics and CVD risk factor control at baseline.

| Characteristic | Controlled CVD risk factors (N=219) | Uncontrolled CVD risk factors (N=252) | p-value |
| --- | --- | --- | --- |
| **Sociodemographic characteristic** |  |  |  |
| Age, mean (SD) | 43.90 (11.25) | 45.91 (10.27) | 0.04 |
| Race |  |  | 0.08 |
| Hispanic | 27.4% | 27.0% |  |
| Non-Hispanic Black | 45.7% | 51.2% |  |
| Non-Hispanic Other | 1.4% | 4.0% |  |
| Non-Hispanic White | 25.6% | 17.9% |  |
| Gender |  |  | 0.42 |
| Female | 9.6% | 11.90% |  |
| Male | 90.4% | 88.10% |  |
| Education |  |  | 0.62 |
| Less than high school | 23.7% | 22.6% |  |
| GED | 22.4% | 19.0% |  |
| High school graduate | 27.4% | 31.0% |  |
| Some college/technical education | 22.4% | 25.0% |  |
| College graduate | 4.1% | 2.4% |  |
| Marital status |  |  | 0.98 |
| Single | 75.3% | 75.4% |  |
| Married | 9.6% | 9.1% |  |
| Separated, widowed divorced | 15.1% | 15.5% |  |
| Average monthly income |  |  | 0.48 |
| No income | 66.8% | 66.1% |  |
| Less than \$500 | 18.4% | 21.0% | |
| \$500-\$999 | 6.0% | 3.2% | |
| \$1000 or more | 8.8% | 9.7% | |
| **Behavioral Factors (post-release)** |  |  |  |
| Smoking | 67.2% | 65.2% | 0.66 |
| Alcohol Use | 4.1% | 4.4% | 0.89 |
| Cocaine Use | 3.2% | 4.4% | 0.51 |
| Physical exercise | 50.7% | 54.4% | 0.42 |
| **Psychosocial factors, mean (SD),** |  |  |  |
| Everyday Discrimination | 15.69 (7.12) | 15.18 (7.08) | 0.44 |
| Perceived Stress Scale | 16.59 (6.89) | 15.20 (7.57) | 0.04 |
| General Self-Efficacy Scale | 33.09(5.24) | 33.02 (5.41) | 0.89 |
| Depression (PHQ-9) | 7.47 (6.10) | 6.78 (5.93) | 0.21 |
| PTSD | 22.22 (13.22) | 18.99 (15.08) | 0.15 |
| Lifetime Cumulative Adversity | 17.72 (6.63) | 15.94 (6.62) | <0.01 |
| **Structural factors** |  |  |  |
| Employed | 12.3% | 15.9% | 0.27 |
| Health Insurance | 92.2% | 94.4% | 0.34 |
| Primary care provider | 46.8% | 46.2% | 0.89 |
| Housing Status |  |  | 0.56 |
| Homeless | 3.2% | 4.8% |  |
| Transitional Home | 80.8% | 75.8% |  |
| Living with Family/Friends | 7.8% | 10.3% |  |
| Rent or Own | 8.2% | 9.1% |  |
| Living in a halfway house | 78.5% | 75.8% | 0.48 |
| **Correctional specific factors/collateral sanctions** |  |  |  |
| Lifetime incarceration |  |  | 0.79 |
| One time | 11.5% | 10.0% |  |
| 2 - 5 times | 41.7% | 45.8% |  |
| 6-10 times | 21.1% | 21.3% |  |
| More than 10 times | 25.7% | 22.9% |  |
| Lifetime incarcerated; months, median (IQR) | 132 (180) | 136 (180) | 0.64 |
| Recent incarceration |  |  | 0.05 |
| Jail | 17.8% | 25.4% |  |
| Prison | 82.2% | 74.6% |  |
| Length of incarceration; months, median (IQR) | 25 (42) | 20 (42) | 0.27 |
| Current Parole | 45.2% | 48.8% | 0.43 |
| Current Probation | 20.1% | 21.0% | 0.8 |
| Solitary confinement | 39.6% | 36.8% | 0.53 |
| Saw health care provider during last incarceration | 68.5% | 73.4% | 0.24 |
| Current SSDI | 5.0% | 3.2% | 0.31 |
| Current Food Stamps | 26.0% | 27.5% | 0.72 |
| Without food for a whole day since release | 20.5% | 19.4% | 0.77 |
| Child Support | 16.4% | 21.8% | 0.14 |
| Barriers to Employment / Licensure based on criminal record | 73.1% | 74.6% | 0.7 |
| Debt / Restitution | 36.1% | 36.1% | 0.99 |
| Barriers to Housing based on criminal record | 7.8% | 8.7% | 0.7 |

A higher proportion of those released from jail had an uncontrolled CVD risk factor compared with those released from prison (62% vs. 51%, p=0.05). Solitary confinement was not associated with CVD risk factor control following release (48% vs. 52%, p=0.53). There were no differences in CVD risk factor control at baseline among those exposed to collateral sanctions of having a criminal record, including those related to employment, housing, or debt. A higher proportion of participants who reported difficulties with paying child support had uncontrolled CVD risk factors compared to those who did not (60% vs. 40%, p=0.14), though the difference was not statistically significant (Table 3).

Average perceived stress score was lower among those with any uncontrolled CVD risk factors compared with those without (15.2 ± 7.6 vs. 16.6 ± 6.9, p=0.04), as was the cumulative adversity score (15.9-± 6.6 vs. 17.7 ± 6.6, p=0.004). There was no difference in scores for general self-efficacy (33.0 ± 5.2 vs. 33.1 ± 5.2, p=0.89), depression as measured by the PHQ-9 (6.8 ± 5.9, 7.5 ± 6.1, p=0.21), or reports of everyday discrimination (15.2 ± 7.1, 15.7 ± 7.2, p =0.44) among those with and without uncontrolled CVD risk factors (Table 3).

In multivariate analysis, after adjusting for participant age and gender, being incarcerated in jail as opposed to prison was associated with an increased odds of having uncontrolled CVD risk factor (AOR 1.59, 95% CI 1.01-2.50). The addition of perceived stress, and life adversity score in the multivariate model did not change the effect size of the association between incarceration in jail on uncontrolled CVD risk factors (AOR 1.62, 95% CI 1.02-2.57) (Figure 1).

**Figure 1.**
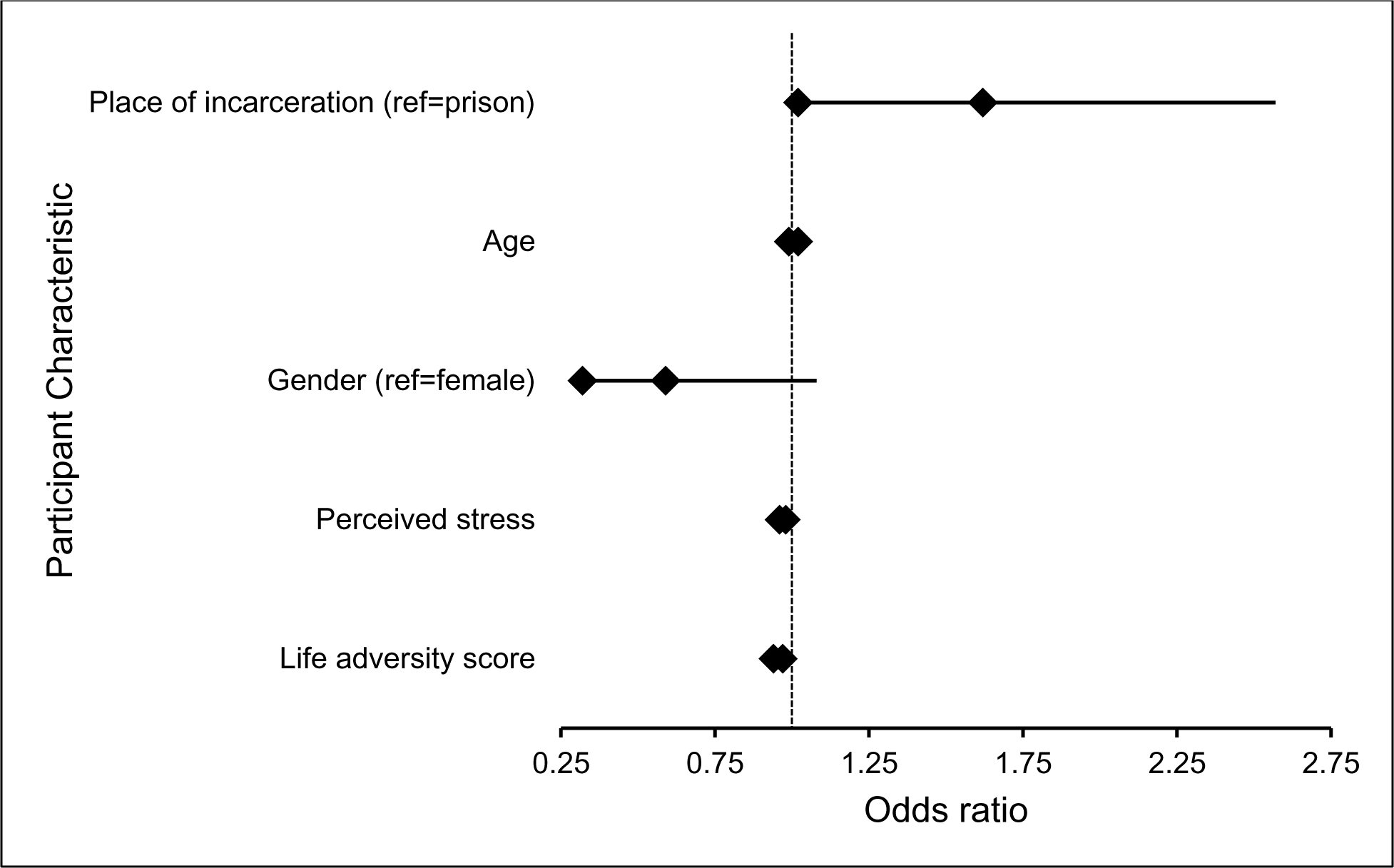
Logistic regression for uncontrolled CVD risk factor at baseline visit.

Our sample’s average LE8 score was 50.64, and total LE8 was not different among those who spent their last incarceration in jail compared to prison (49.85 vs 50.86). Among the eight domains of LE8, blood pressure (p=0.02) and smoking (p<0.001) were significantly worse among those incarcerated in jail compared with those incarcerated in prison, while diet was better among those who had been incarcerated in jail compared with prison (Table 4).

**Table 4:**
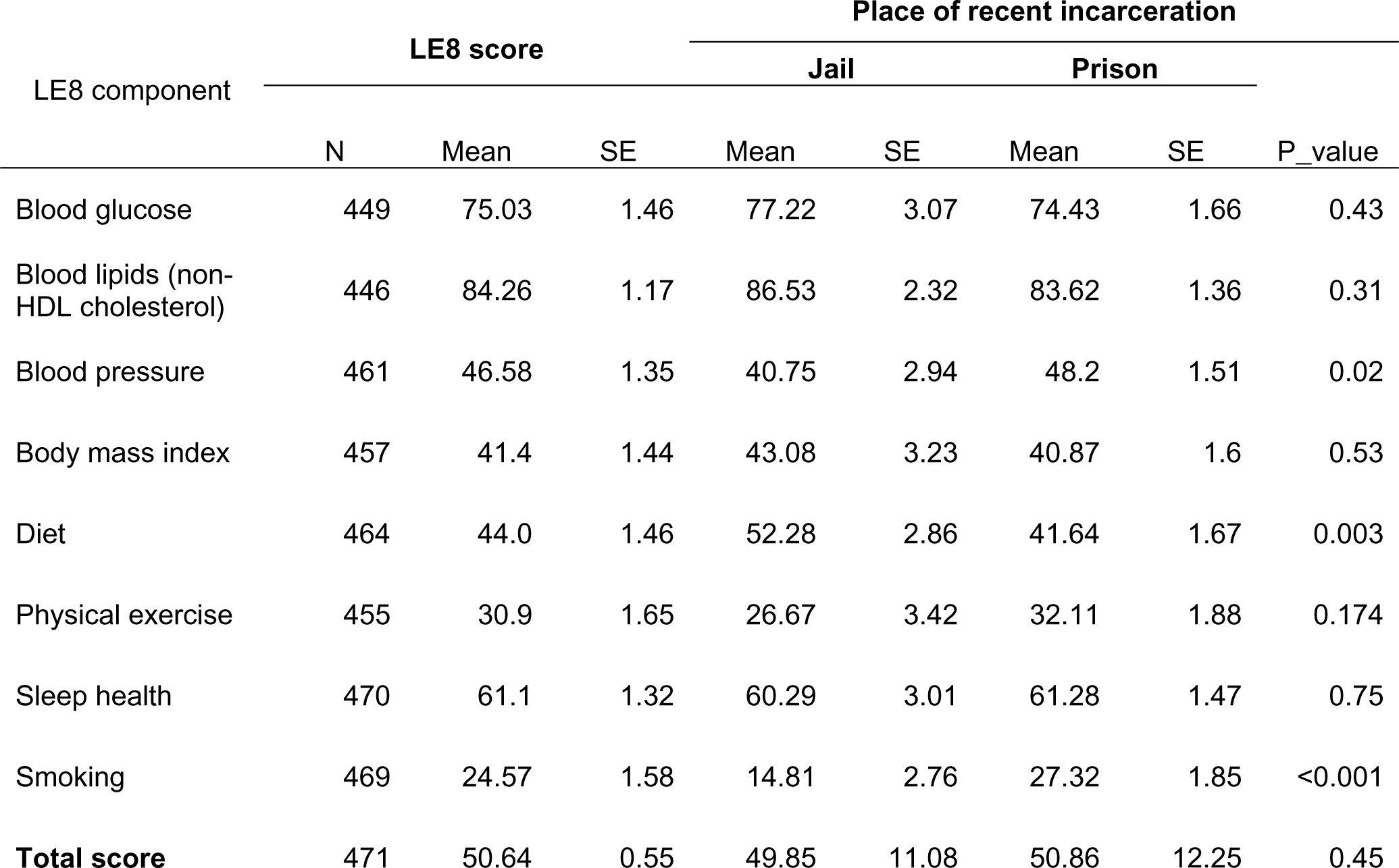
Life’s Essential 8 scores stratified by Place of Last Incarceration.

| LE8 component | LE8 score |  |  | Place of recent incarceration |  |  |  |  |
| --- | --- | --- | --- | --- | --- | --- | --- | --- |
|  |  |  |  | Jail |  | Prison |  | P_value |
|  | N | Mean | SE | Mean | SE | Mean | SE |  |
| Blood glucose | 449 | 75.03 | 1.46 | 77.22 | 3.07 | 74.43 | 1.66 | 0.43 |
| Blood lipids (non-HDL cholesterol) | 446 | 84.26 | 1.17 | 86.53 | 2.32 | 83.62 | 1.36 | 0.31 |
| Blood pressure | 461 | 46.58 | 1.35 | 40.75 | 2.94 | 48.2 | 1.51 | 0.02 |
| Body mass index | 457 | 41.4 | 1.44 | 43.08 | 3.23 | 40.87 | 1.6 | 0.53 |
| Diet | 464 | 44.0 | 1.46 | 52.28 | 2.86 | 41.64 | 1.67 | 0.003 |
| Physical exercise | 455 | 30.9 | 1.65 | 26.67 | 3.42 | 32.11 | 1.88 | 0.174 |
| Sleep health | 470 | 61.1 | 1.32 | 60.29 | 3.01 | 61.28 | 1.47 | 0.75 |
| Smoking | 469 | 24.57 | 1.58 | 14.81 | 2.76 | 27.32 | 1.85 | <0.001 |
| **Total score** | 471 | 50.64 | 0.55 | 49.85 | 11.08 | 50.86 | 12.25 | 0.45 |

## Discussion

Release from jail, compared with prison, is significantly associated with having an uncontrolled CVD risk factor, amongst those with underlying hypertension, diabetes, hyperlipidemia, or obesity. This finding has bearing on population health, given that 7 million individuals move through jails yearly in the United States, and more than 62% have a known cardiovascular risk factor.^37^ It is plausible that the disruption to the life by being incarcerated is what disrupts CVD risk factor management. Even one day in jail can lead to the loss of employment, housing, and healthcare. Further, medications and devices available in the community are often different or unavailable in carceral facilities and thus can lead to worsening of CVD risk factor control.^38^

Surprisingly, while the vast majority of participants report having health insurance upon release, less than half of these individuals report having a primary care physician upon release, supporting other research showing that enrollment in Medicaid is necessary though not sufficient to engage individuals just released from the carceral system into primary care. Also, though those released from jail have higher rates of uncontrolled CVD risk factors, the rates of those in prison were high, and there remains a need to address risk factors in the entire population of those who have experienced incarceration.

These data are relevant because jails and prisons have been some of the only places in the US where federal Medicaid and Medicare funds cannot be used to pay for health care services due to the Medicaid Inmate Exclusion Policy.^39^ But the design and implementation of the Centers of Medicare & Medicaid Service’s new Medicaid 1115 Reentry Waiver may allow state Medicaid programs for the first time to cover services that address chronic health conditions like CVD and its risk factors as individuals transition from carceral systems back to the community. With this historic change of policy, Medicaid can now be used to cover services for people who are incarcerated and eligible for Medicaid and returning home to their communities within 90 days. Already, two states have 1115 waivers approved, with at least 20 more applications pending.^40^ Our data indicate that to improve the disproportionately high hospitalization and death rates from CVD immediately upon release from correctional systems, jails should be included as targets of intervention.

Furthermore, our results demonstrate that focusing on hypertension control and smoking cessation are important areas to prioritize for quality-of-care improvement – as they are both highly prevalent conditions that cause significant morbidity following release. A recent feasibility trial in an urban jail found that nicotine replacement therapy was feasible to initiate and maintain following release, though there were no differences in abstinence rates between the groups that received standard behavioral health education compared with nicotine replacement therapy and counseling.^41^ Future work needs to be conducted on how best to improve the transition of healthcare in and out of jail, which are usually short stay facilities, to the community for CVD risk factors and may also require examining how incarceration (movement from the community to the jail) affects these outcomes.

We found that people with uncontrolled CVD risk factors had lower rates of psychosocial stress and lifetime cumulative adversity than those with controlled CVD risk factors. These data conflict with large studies of non-incarcerated people that find that psychosocial factors and lifetime cumulative adversity are associated with worse uncontrolled CVD risk factors and incident CVD.^42,43^ One plausible explanation noted in the literature is that experience with previous stressors creates resilience in individuals and later produces less stress even when life is out of control. For instance, racially minoritized populations who have consistently lived under economic and social adversities have reported lower psychosocial stress rates than their White counterparts.^44^ Also, several studies have started examining the connection between biological measures of stress and exposure to incarceration at the individual level, which provides an opportunity to evaluate the physiologic manifestations of stress, even if they are not recognized at the same psychological level. For example, a recent study using data from the National Longitudinal Adolescent Health Survey, found a relationship between incarceration lengths of greater than a year and elevated CRP levels, even after adjusting for sociodemographic and clinical factors.^45^ Another study found incarceration exposure predicted epigenetic accelerated aging; more frequent incarceration episodes were associated with accelerated aging as measured by GrimAge in a dose-dependent manner,^46^ even after adjustment for neighborhood factors.^47^ These findings suggest exposure to incarceration may trigger a response that affects a biological stress signature of physiological deterioration, independent of perceived psychosocial stress. Longitudinal data with direct biological measures of stress will be important to examine the temporal relationship between psychosocial stress, lifetime cumulative adversity, and CVD risk factors.

This study has several limitations. For one, no conclusions about cause and effect can be derived from this study due to its cross-sectional design. For example, worse CVD risk factor outcomes may contribute to incarceration in jails instead of prison; individuals with more advanced disease and poor functional health status may be more likely to be sentenced to shorter sentences, though we adjusted for age in multivariate analysis. These effect-cause relationships deserve further exploration in future longitudinal analyses. This study was conducted in Connecticut among people just released from carceral facilities who largely have access to health insurance immediately post release. Further studies would need to explore the association between incarceration-specific risk factors and uncontrolled CVD risk factors in other localities, especially in the South, where the use of solitary confinement and collateral sanctions following release is far higher. Additionally, given that Connecticut has a unified correctional system with the same health system and electronic health record in prisons, this study is biased toward finding less of a difference between those incarcerated in jail and prison, and thus our findings may underestimate this difference in other states. The length of incarceration jail is longer than is typical most jail stays in the country and may not be reflective of all carceral facilities, as presentencing during COVID-19 substantially lengthened in Connecticut and throughout the country. Lastly, this study recruited during the COVID-19 pandemic when people in jails, prisons, and halfway houses were subject to tremendous stressors given facility-wide lockdowns, quarantine and isolation policies, and high COVID-19 morbidity and mortality rates in correctional facilities. These conditions likely affected managing CVD risk factors control in carceral systems and following release.

## Conclusion

In a statewide study of a unified correctional system, having been released from a jail, a pre-sentencing and shorter-sentence carceral facility, was associated with higher rates of uncontrolled CVD risk factors than being released from prison. Plausible explanations include disruptions in employment, healthcare, and family supports and poor transition of care from the community into jail and back. Longitudinal studies designed to better understand the relationships between exposure to mass incarceration and CVD risk factors are needed to inform specific interventions to achieve cardiovascular health equity. Medicaid Reentry Section 1115 Demonstration projects provide an opportunity to specifically focus on interventions to improve cardiovascular health and should include a focus on those being released from jails.

## Funding

This work is supported by the National Heart, Lung, and Blood Institute grant R01 HL137696, awarded to Dr. Emily Wang.

## Role of the Funder

The National Heart, Lung, and Blood Institute had no role in the design and conduct of the study; management, analysis, and interpretation of the data; manuscript preparation, and decision to submit the manuscript for publication.

## Author Disclosures

Dr Harlan Krumholz reported receiving expenses and/or personal fees from Element Science, Eyedentify, and F-Prime, and is a co-founder of Hugo Health, Refactor Health, and Ensight-AI. He is associated with contracts, through Yale New Haven Hospital, from the Centers for Medicare & Medicaid Services and through Yale University from Janssen, Johnson & Johnson Consumer, and Pfizer, outside the submitted work. No other disclosures were reported.

## Author contributions

JAA: Methodology, Data curation, Formal analysis, Writing-Original draft preparation, Writing-Reviewing and Editing. LBP: Writing- Original draft preparation, Writing- Reviewing and Editing. BR: Writing- Reviewing and Editing. NH: Writing- Reviewing and Editing. JEE: Writing- Reviewing and Editing. HL: Writing- Reviewing and Editing. KBD: Writing- Reviewing and Editing. HK: Writing- Reviewing and Editing. EAW: Conceptualization, Methodology, Funding acquisition, Writing- Original draft preparation, Writing- Reviewing and Editing, Supervision.

## Disclaimer

The analysis, interpretation or conclusions drawn from these data are the responsibility of the authors and do not represent the views of neither the National Heart, Lung, and Blood Institute nor the United States Department of Health and Human Services or any of its affiliates. The authors assume full responsibility for all analyses, interpretations, and conclusions.

## Prior presentation

EPI | Lifestyle 2024 Scientific Sessions; Chicago March 18-21, 2024.

## Acknowledgement

We are grateful and acknowledge the efforts of the study team members who consented and collected participant data (Reynold Jules, Bernice Kwarteng, Cerella Criag, Marisol Foumakoye and Edgar Jones). We also acknowledge outreach efforts by Monya Saunders and Tino Negron to identify potential participants, and the collaboration of Intercommunity Health care.

## Data availability statement

The data that support the findings of this study are available from the principal investigator (EAW) upon reasonable request.

## Notes

### Clinical Trial

Not applicable

### Author Declarations

The Yale Human Investigation Committee and the Office of Human Research Protections approved this study.

